# Anaemic women are more at risk of injectable contraceptive discontinuation due to side-effects in Ethiopia

**DOI:** 10.1101/2020.10.28.20221523

**Authors:** Rose Stevens, Blandine Malbos, Eshetu Gurmu, Jérémie Riou, Alexandra Alvergne

## Abstract

**Introduction:** This paper investigates the importance of women’s physiological condition for predicting the risk of discontinuation due to side-effects of the injectable contraceptive in Ethiopia, where side-effects account for around 20% of all discontinuations.

**Methods:** Contraceptive calendar data from the 2016 Ethiopian Demographic and Health Survey were analysed. Women aged 15-49 who had initiated the injectable contraceptive in the two years prior to interview were included in the analysis (N=1,513). After checking for reverse causality, the associations between physiological risk factors and discontinuation of the injectable contraceptive due to either side-effects (DSE) or other reasons (DOR) were estimated using multivariate Cox proportional regression analyses.

**Results:** In 2016, 10% of women had initiated the injectable in the last two years, and 1 in 4 had discontinued use by the time of the interview. Of these, 1 in 5 discontinued due to side-effects. Women with anaemia were at twice the risk of DSE compared with non-anaemic women, while anaemia status was not associated with DOR. The association between anaemia and experience of side-effects is likely driven by iron-deficiency anaemia, as having taken iron supplements during last pregnancy is found to decrease the risk of DSE. Sociocultural factors including religion, wealth and relationship status were significant predictors for DOR, but not for DSE.

**Conclusion:** Accounting for diversity in physiological condition is key for understanding contraceptive discontinuation due to side-effects. To reduce side-effects and thereby unmet need for contraception, family planning programs may benefit from providing an integrated service package addressing anaemia as well as supplying hormonal contraception.

## Introduction

Globally, over a third of women discontinue modern contraception within a year of initiating use and among those who discontinue whilst still in need of contraception, they primarily do so due to the experience of negative side-effects.^1^ Hormonal contraceptive side-effects in particular, such as irregular bleeding, headaches, and nausea, take a heavy toll on women’s lives. Yet, why some women experience more side-effects than others has attracted little research to date. Clinical trials are focused on overall side-effects burden rather than interindividual variation^2^ and public health research concentrates efforts on dispelling misconceptions and myths.^3^ There has been comparatively little research into potential biological explanations for patterns of diversity in the experience of side-effects, despite suggestions that women in poorer physiological condition may experience a greater burden of side-effects.^4^

Most scholarly arguments explaining why women discontinue using modern contraceptives whilst in need have focused on sociocultural factors which impact contraceptive knowledge, attitudes and service barriers.^5^ The experience of and fear of side-effects are often conflated, with the underlying assumption that many fears are not grounded in real experience and can be dispelled through counselling.^3^ This information-deficit model biases the analytical strategy of current efforts aimed at understanding contraceptive discontinuation. For instance, studies do not usually disaggregate discontinuation by reason when assessing risk factors^6–9^. Further, the focus is on sociocultural causes such as religion, age, education, and wealth, rather than explicit measures of physiological condition and poor health. Studies also do not usually disaggregate by contraceptive method, or by hormonal vs. non-hormonal methods, which may preclude the identification of causal pathways between physiology and side-effects. A new model, taking into account a potential biological aetiology, is needed to explain interindividual variation in the experience of contraceptive side-effects.

A biological perspective suggests that side-effects arise from the interaction of synthetic hormones with women’s physiology. The occurrence of side-effects will be dependent on the dose of the drug, the time course over which it acts, and other susceptibility factors. The type of side-effects experienced depends on the concentration of synthetic hormones at targeted and/or non-targeted sites of action, as well as disturbances of natural reproductive hormone levels. Women who experience the greatest suppression of natural reproductive hormones after initiating hormonal contraception are those with the lowest levels of natural reproductive hormones prior to use.^10^ This greater suppression of natural hormones may cause them to experience more severe side-effects, such as symptoms associated with natural hormone deficiency (e.g. mood swings, headaches, irregular bleeding). If women with lower levels of natural reproductive hormones experience more intense/frequent side-effects,^4^ then diversity in women’s reproductive physiology could explain variation in discontinuation due to side-effects.

Anthropological studies have shown that the lowest concentrations of reproductive hormones are found among women with poor physiological condition due to energetic and immune stress^11,12^, associated with indicators such as high workloads, long times to fetch water, high inflammation or low body size and fat stores. Another indicator of poor somatic condition is anaemia, which might indicate low natural hormone levels. Anaemia can result from environmental and somatic stressors, such as iron deficiency, due to low dietary iron or depletion from pregnancy, and/or from infection. In turn, anaemia reduces the function of the granulosa cells, which make up the follicle surrounding a maturing egg^13^. This may lead the ovaries to produce lower levels of reproductive hormones. Though there is a lack of clinical evidence for the relationship between markers of physiological condition and side-effects, women in qualitative studies regularly cite harsh lifestyle factors and poor diets as predisposing individuals to experiencing contraceptive side-effects^14,15^. This relationship has yet to be investigated quantitatively.

This paper seeks to investigate the association between women’s physiological condition and discontinuation due to side-effects (DSE) in the Ethiopian 2016 Demographic and Health Survey (DHS).^16^ Not all variables which may be associated with reproductive hormone levels were available in this dataset, but it provides a high-quality large-scale dataset with detailed contraceptive information and relevant physiological factors. Rates of DSE are high in Ethiopia (about 1 in 5 women reported “side-effects” as the main reason for discontinuation) despite an innovative and extended community-based Family Planning Program. We focus our analysis on the 3-month injectable contraceptive, as it is the most prevalent method in Ethiopia (63%)^16^ and Sub-Saharan Africa (47%).^17^ It is also consistently is associated with the highest rates of DSE,^18^ and allows us to eliminate any confounding by method type. To evaluate the role of physiological condition in increasing risk of DSE specifically, as opposed to any form of discontinuation, we compared the association of markers of physiological condition with both discontinuation due to side-effects (DSE) and discontinuation due to other reasons (DOR). We predicted that measures of physiological stress (low BMI, agricultural occupation, longer time to fetch water, being anaemic, lack of iron supplementation, recent birth prior to injectable initiation) would increase the risk of DSE, but not of DOR. This knowledge may help devise new solutions tailored to women’s bodies to reduce side-effects and improve continuation.

## Methods

### Data and participants

#### The DHS Program

This study focuses on the Ethiopian 2016 DHS survey, which was implemented by the Central Statistical Agency (CSA) from January to June 2016.^16^ Procedures and questionnaires for standard DHS surveys have been approved by the ICF Institutional Review Board (IRB) and the specific Ethiopian protocol by IRBs in Ethiopia. A total of 16,650 households were successfully interviewed, with a response rate of 98% of occupied households.^16^ The DHS survey is cross-sectional in nature and it uses a two-stage cluster sampling approach. Sample weights were employed in order to obtain population representative results.

#### Data Collection Methods

An informed consent statement was read to the respondents before the interview. Detailed information on lifestyle, health, contraceptive and reproductive histories were collected for 15,683 women aged 15-49. Histories were collected using the ‘reproductive-contraceptive calendar’. This is a month-by-month history of the last 5-6 years preceding the interview of a woman’s key reproductive events. This method of collecting contraceptive histories has been shown to be of better quality than standard questionnaire methods.^19^ Only one reason per discontinuation event is recorded, precluding any analysis of multifactorial discontinuation decisions. A drop of blood taken from a finger prick was analysed using a HemoCue to measure haemoglobin levels. Haemoglobin was then adjusted according to WHO recommendations for cigarette smoking and altitude.^20^ These adjustments may not be appropriate for certain East African ethnicities,^21^ but we retained them due to lack of research on haemoglobin adaptations across ethnicities and to allow for comparability with other studies.

#### Inclusion Criteria

Only women who initiated injectable contraception within two years prior to the interview were included to reduce recall bias and eliminate left censoring. This ensured that biomarker data taken at interview, such as haemoglobin and BMI, were taken a maximum of two years since contraceptive initiation, a period over which they were assumed to be relatively stable. Haemoglobin trajectories have been shown to be consistent during this timeframe.^22^ Only the first episode of injectable use in the last two years was included in the analysis. We excluded women who were pregnant at the time of the interview and women who answered ‘Other’ for religion due to the small sample size (n=8). A total of 1,513 women aged 15-49 were included in the final analysis (Figure 1).

**Figure 1:**
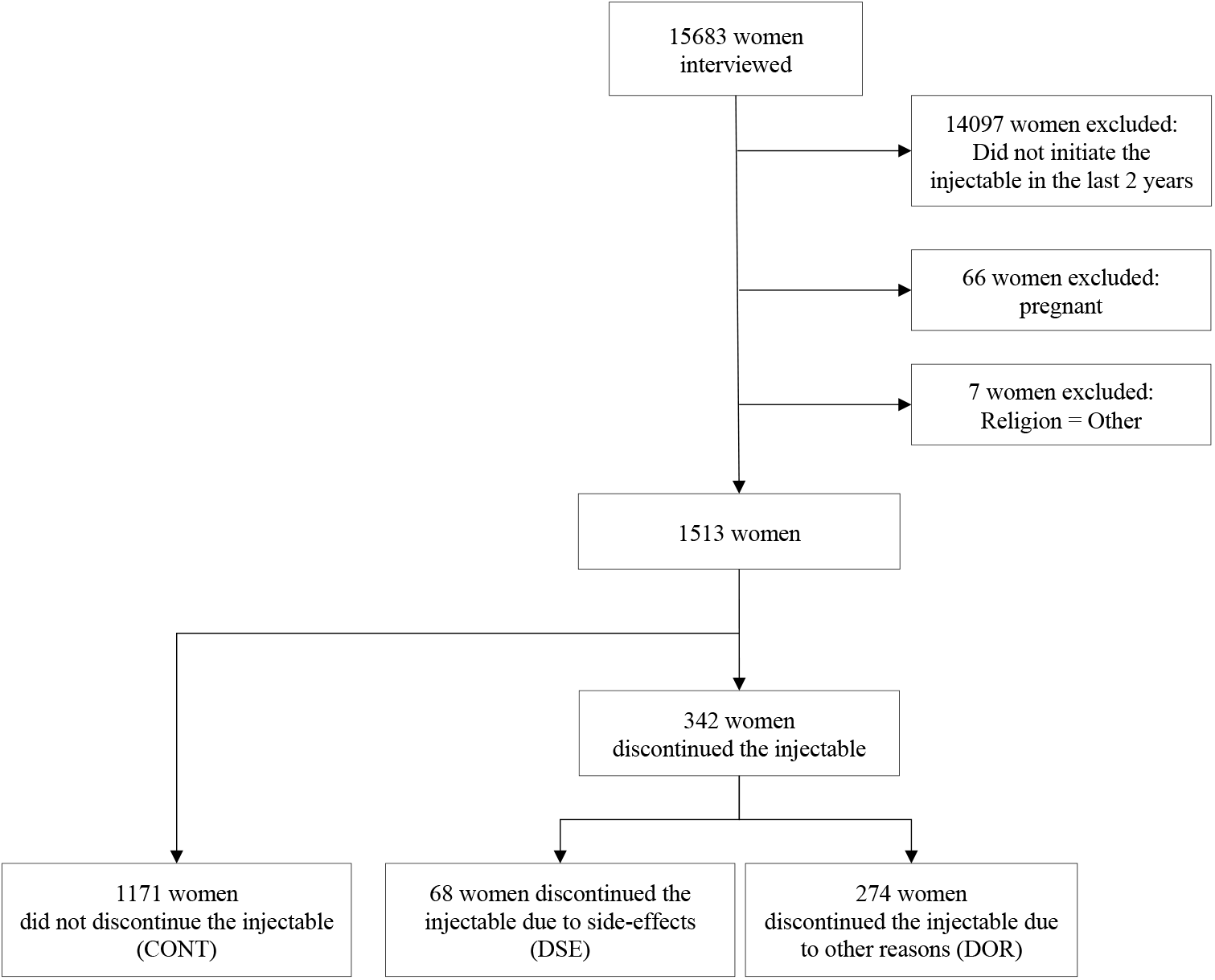
Sample flow chart showing inclusion criteria.

### Variables

#### Outcome variables

Our primary outcome variable was time from initiation to discontinuation due to side-effects (DSE). Our secondary outcome variable was time to discontinuation due to other reasons other than side-effects (DOR).

#### Independent variables

We extracted standard sociodemographic variables and known risk factors for discontinuation, such as age (15-24, 25-34, 35-49), education (no, primary, secondary+), residence de facto (urban, rural), ethnicity (Amhara, Oromo, Tigrie, Others; 80 levels brought down to 4 based on prevalence), parity (0, 1-2, 3+), religion (Muslim, Orthodox Christian, Catholic and Protestant), household wealth (40% poorest, 20% middle, 40% richest; derived from 5 quintiles, based on a score derived from various consumer goods owned and housing characteristics), and relationship status (Yes (married or cohabiting), No).

In order to evaluate the importance of physiological condition, we extracted the variables height (in cm; using a Shorr measuring board) and weight (in kg; using a SECA scale) to compute BMI (Underweight <18.5, Normal: 18.5-24.9, Overweight ≥25.0), time to fetch water (≤20 min, <20min), occupation (agricultural/non-agricultural/no work), anaemia (Yes, No; classified as anaemic if adjusted haemoglobin was <12g/dl), taken iron supplementation (Yes, No; only collected for supplementation during last pregnancy in last 5 years) and time from most recent birth to injectable initiation (0-2 months, 3-12 months, 13-24 months, >24 months, No births in the last 5 years).

### Statistical analysis

#### Reverse causality

An analysis based on weighting propensity score using an Inverse Probability Weighting (IPW)^23,24^ was performed to evaluate the causal impact of currently taking injectables on anaemia and BMI. This weighting allows two groups (current injectable use (minimum 3 months) versus no contraceptive method or pregnancy in the last 24 months), strictly comparable on all the variables included in the survival analysis to follow. The balance of these different variables, potential confounding factors, is summarized in Figure S1. This strategy allows the ability to approach the design of experimental studies and envisage a causal effect of the injectable on the variables of interest.

#### Survival Analysis

We tested for exposure variables associated with time to discontinuation due to side-effects (DSE). Our secondary analysis tested for exposure variables associated with the secondary endpoint, i.e. time to discontinuation due to other reasons (excluding side-effects) (DOR). This secondary analysis allows comparison of results to ensure any significant effect found is specific to side-effects, as opposed to simply discontinuation. The original analysis strategy for this research and predicted results were pre-registered with the Open Science Framework before data were observed (registration can be found at https://osf.io/276gt). Some changes were made from the original analysis plan, but in essence, it remained the same.

Multivariate cox regression analyses were performed using the free R software^25^ to compare the impact of exposure variables on both time to DSE and time to DOR. Women were right-censored either at the time of the survey or earlier if they discontinued for other reasons. The unit of analysis is an episode (i.e. a month) during which women are ‘at risk’ of discontinuing. The hazard of an event (i.e. DSE or DOR) is predicted as a function of the duration of injectable use.

Exposure variables associated with either DSE or DOR to p<0.2 in univariate Cox regression analyses were included in the multivariate Cox regression analysis together with known confounders. To obtain a parsimonious final model, a manual step-down variable selection coupled with a study of the Akaike criterion was performed for each regression model. A check for collinearity between the explanatory variables was performed using the variance inflation factor (VIF). Hazard-ratio (HR), 95% confidence interval (CI) and p-value were used to determine if effects were significant. Proportionality was checked using scaled Schonfeld residuals, both by plotting these against time and testing formally for non-proportionality.

All analyses were performed at the Type-I error rate (alpha) of 5 percent. All tests were two-sided. The packages ‘survival”, ‘ggplot2’, ‘WeigthIt’, ‘survey’, ‘missForest” were used^26,27^. Due to the prioritization of objectives, no correction for multiple testing was necessary. No data imputation was made. An analysis of missing data has been performed (Figure S2, Figure S3). If these are MNAR (Missing Not at Random), then our conclusions will be affected and discussed.

## Results

### Descriptive statistics

Among the 1,513 women aged 15-49 who initiated the injectable the two years preceding the DHS interview, 25.7% discontinued the injectable. Of those who discontinued, 19.9% women reported side-effects as their reason for discontinuation, second only to wanting to become pregnant (21.9%). Our analysis is minimally affected by missing data (only 1.7% of all data missing), which are not informative (Figure S2, Figure S3). Differences in subjects’ characteristics between groups were analysed using the Fisher exact test for categorical variables. Descriptive analysis was stratified according to the outcome of interest (Table 1). Women in the DSE group were more likely to be anaemic and more likely to want another child as compared to the non-DSE group. Women who DSE were less likely to have taken iron supplements with their last pregnancy and less likely to have had a baby in the last 2 months. By contrast, women in the DOR group were no different in respect to anaemia compared to the non-DOR group, but were more likely to be underweight, younger, have fewer children, and want another child (Table 1).

**Table 1:** Weighted descriptive table of the sample for all variables included in the analysis. DSE: Discontinuation due to side-effects. DOR: Discontinuation due to other reasons. P-value shows results of Fisher exact test. N weighted = 1925. N unweighted = 1513.

|  | TOTAL | Comparison between NON-DSE and DSE |  |  | Comparison between NON-DOR and DOR |  |  |
| --- | --- | --- | --- | --- | --- | --- | --- |
|  | n (%) | NON-DSE<br>n (%) | DSE<br>n (%) | p-value | NON-DOR<br>n (%) | DOR<br>n (%) | p-value |
| **Age group** |  |  |  | 0.096 |  |  | >0.001 |
| [15.24] | 655 (34%) | 637 (35%) | 18 (23%) |  | 517 (32%) | 138 (45%) |  |
| [25.34] | 880 (46%) | 837 (45%) | 43 (54%) |  | 763 (47%) | 116 (38%) |  |
| [35.49] | 390 (20%) | 372 (20%) | 18 (23%) |  | 336 (21%) | 53 (17%) |  |
| **Occupation** |  |  |  | 0.111 |  |  | 0.151 |
| Agricultural work | 434 (23%) | 409 (23%) | 25 (33%) |  | 357 (23%) | 77 (25%) |  |
| No working | 953 (51%) | 919 (51%) | 34 (45%) |  | 815 (51%) | 138 (45%) |  |
| Non-agricultural work | 499 (26%) | 482 (27%) | 17 (22%) |  | 411 (26%) | 89 (29%) |  |
| **Area** |  |  |  | 0.670 |  |  | 0.521 |
| Urban | 356 (19%) | 343 (19%) | 13 (17%) |  | 1321 (82%) | 247 (80%) |  |
| Rural | 1568 (81%) | 1503 (81%) | 65 (83%) |  | 295 (18%) | 61 (20%) |  |
| **Parity** |  |  |  | 0.106 |  |  | >0.001 |
| 0 | 253 (13%) | 248 (13%) | 5 (6%) |  | 189 (12%) | 65 (21%) |  |
| [1-2] | 711 (37%) | 685 (37%) | 26 (34%) |  | 574 (34%) | 137 (44%) |  |
| 3+ | 959 (50%) | 913 (49%) | 46 (60%) |  | 853 (53%) | 106 (34%) |  |
| **Anaemia** |  |  |  | 0.007 |  |  | 0.427 |
| No | 1514 (81%) | 1462 (82%) | 52 (69%) |  | 1277 (81%) | 237 (80%) |  |
| Yes | 351 (19%) | 328 (18%) | 23 (31%) |  | 290 (19%) | 61 (20%) |  |
| **BMI (kg/m<sup>2</sup>)** |  |  |  | 0.852 |  |  | 0.002 |
| <18.5 | 363 (19%) | 350 (19%) | 13 (17%) |  | 282 (18%) | 80 (27%) |  |
| [18.5;25[ | 1405 (75%) | 1347 (75%) | 58 (76%) |  | 1200 (76%) | 205 (68%) |  |
| ≥ 25 | 108 (6%) | 103 (6%) | 5 (7%) |  | 93 (6%) | 16 (5%) |  |
| **Wealth index** |  |  |  | 0.175 |  |  | 0.007 |
| 40% poorest | 610 (32%) | 582 (32%) | 28 (36%) |  | 489 (30%) | 121 (40%) |  |
| 40-60% | 438 (23%) | 427 (23%) | 11 (14%) |  | 375 (23%) | 64 (21%) |  |
| 40% richest | 876 (46%) | 837 (45%) | 39 (50%) |  | 752 (47%) | 123 (40%) |  |
| **Education** |  |  |  | 0.618 |  |  | 0.928 |
| No education | 981 (51%) | 938 (51%) | 43 (56%) |  | 822 (51%) | 159 (52%) |  |
| Primary | 672 (35%) | 649 (35%) | 23 (30%) |  | 565 (35%) | 107 (35%) |  |
| Secondary+ | 270 (14%) | 259 (14%) | 11 (14%) |  | 228 (14%) | 41 (13%) |  |
| **Religion** |  |  |  | 0.620 |  |  | >0.001 |
| Protestant + Catholic | 497 (26%) | 480 (26%) | 17 (22%) |  | 464 (29%) | 33 (11%) |  |
| Muslim | 463 (24%) | 445 (24%) | 18 (23%) |  | 389 (24%) | 74 (24%) |  |
| Orthodox | 964 (50%) | 921 (50%) | 43 (55%) |  | 763 (47%) | 201 (65%) |  |
| **Ethnicity** |  |  |  | 0.053 |  |  | >0.001 |
| Amhara | 697 (36%) | 675 (37%) | 22 (29%) |  | 552 (34%) | 145 (47%) |  |
| Oromo | 559 (29%) | 534 (29%) | 25 (32%) |  | 486 (30%) | 73 (24%) |  |
| Tigrie | 172 (9%) | 159 (9%) | 13 (17%) |  | 129 (8%) | 43 (14%) |  |
| Others | 495 (26%) | 478 (26%) | 17 (22%) |  | 448 (28%) | 47 (15%) |  |
| **Desire for a child** |  |  |  | >0.001 |  |  | >0.001 |
| No | 742 (39%) | 726 (39%) | 16 (21%) |  | 650 (40%) | 92 (30%) |  |
| Wants after 2+ years | 943 (49%) | 902 (49%) | 41 (53%) |  | 785 (49%) | 158 (51%) |  |
| Wants unsure tinning | 56 (3%) | 48 (3%) | 8 (10%) |  | 43 (3%) | 12 (4%) |  |
| Wants within 2 years | 183 (10%) | 170 (9%) | 13 (17%) |  | 137 (9%) | 46 (15%) |  |
| **Time to get water** |  |  |  | 0.226 |  |  | 0.072 |
| ≤ 20min | 1013 (55%) | 979 (55%) | 34 (48%) |  | 837 (54%) | 176 (60%) |  |
| >20min | 832 (45%) | 795 (45%) | 37 (52%) |  | 714 (46%) | 119 (40%) |  |
| **Relationship** |  |  |  | 0.716 |  |  | >0.001 |
| No | 170 (9%) | 164 (9%) | 6 (8%) |  | 1519 (94%) | 234 (76%) |  |
| Yes | 1754 (91%) | 1682 (91%) | 72 (92%) |  | 97 (6%) | 73 (24%) |  |
| **Time since last birth to initiation** |  |  |  | 0.003 |  |  | >0.001 |
| 0-2 months | 556 (29%) | 544 (30%) | 12 (16%) |  | 485 (30%) | 71 (24%) |  |
| 3-12 months | 430 (23%) | 406 (22%) | 24 (32%) |  | 364 (23%) | 66 (23%) |  |
| 13-24 months | 315 (17%) | 294 (16%) | 21 (28%) |  | 275 (17%) | 40 (14%) |  |
| > 24 months | 211 (11%) | 207 (11%) | 4 (5%) |  | 187 (12%) | 25 (9%) |  |
| No births in last 5yrs | 394 (21%) | 380 (21%) | 14 (19%) |  | 303 (19%) | 91 (31%) |  |
| **Iron supplementation with last pregnancy** |  |  |  | 0.013 |  |  | 0.003 |
| Not taken | 1080 (59%) | 1029 (58%) | 51 (73%) |  | 908 (57%) | 173 (67%) |  |
| Taken | 764 (41%) | 745 (42%) | 19 (27%) |  | 678 (43%) | 86 (33%) |  |

#### Risk of discontinuation due to side-effects (DSE)

To investigate the role of factors that are specific to DSE, as compared to DOR or continuation, we compared DSE and non-DSE groups. Anaemic women were at over double the odds of DSE than non-anaemic women (HR=2.38, CI= [1.41-4.00], p=0.001) (Table 2, Figure 2). Further, women who had given birth either 3-12 months or 13-24 months prior to injectable initiation were at greater risk of DSE as compared with women who had given birth over 24 months ago, and those who had taken iron supplements were at reduced risk of DSE compared to those that had not (HR=0.54, CI= [0.31-0.96], p=0.036). Finally, as compared to underweight women, overweight women were at greater odds of DSE (HR=3.90, CI= [1.24-12.23], p=0.020).

**Table 2:**
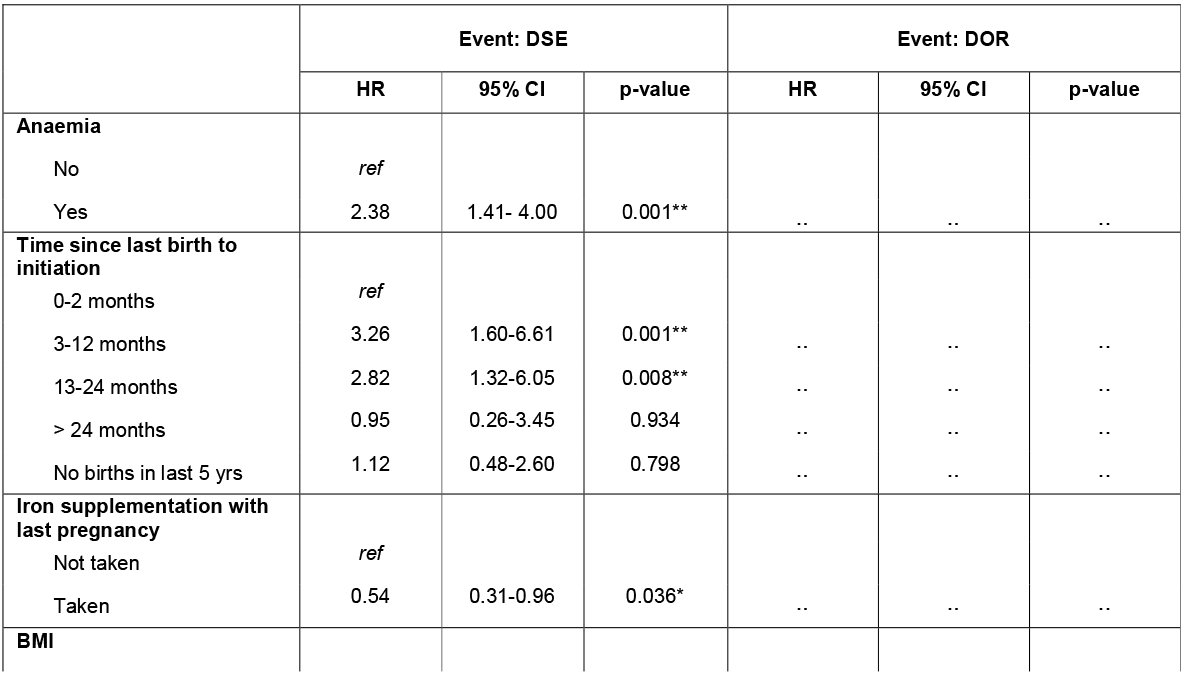
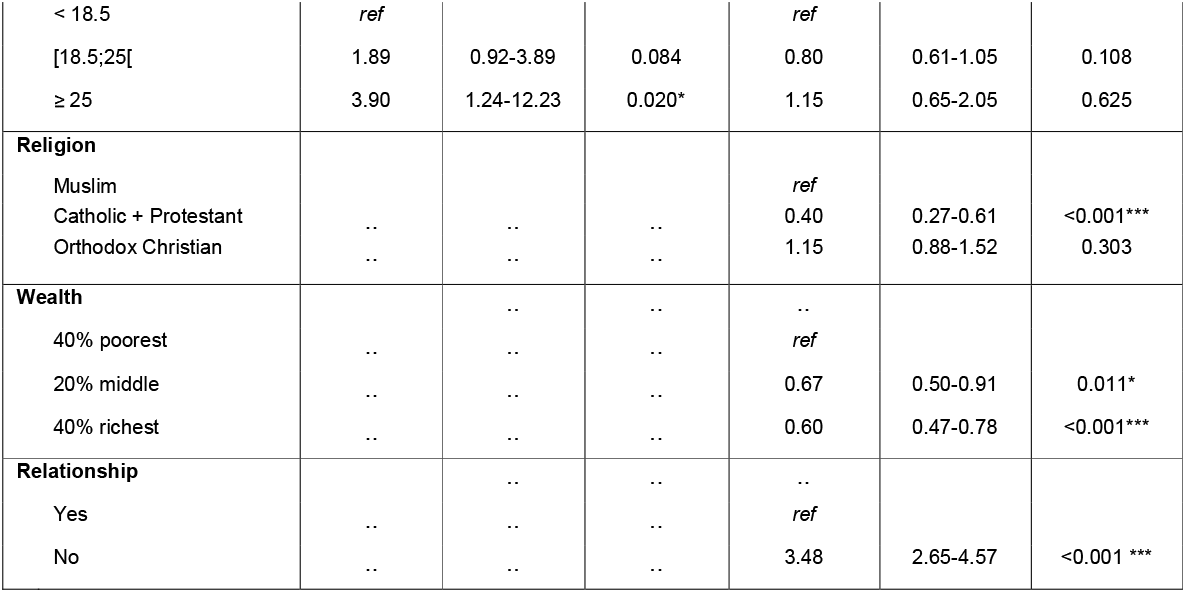
Model outputs showing adjusted hazard ratios from Cox models. *p<0.05, **p<0.01, ***p<0.001.

**Figure 2:**
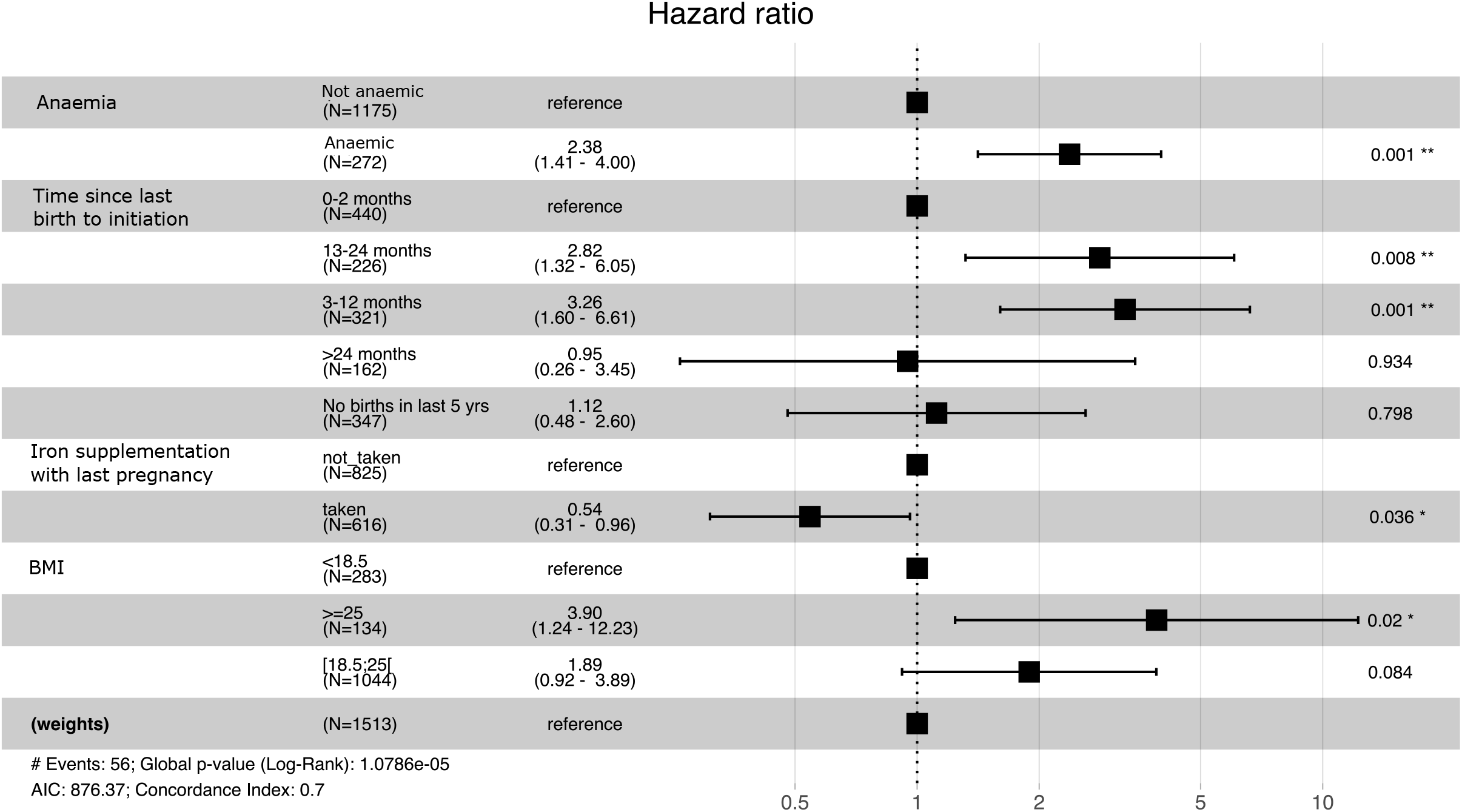
Adjusted hazard ratios and 95% CI for each risk factor for discontinuation due to side-effects (DSE)

In our reverse causality analysis, injectable use was associated with a lower risk of being anaemic at interview and a higher risk of high BMI at interview. Thus, those who were anaemic at the time of interview and DSE are unlikely to have become anaemic due to injectable use, strengthening our assumption that women who DSE and were anaemic were likely anaemic upon injectable initiation.

#### Risk of discontinuation due to other reasons (DOR)

To investigate if physiological risk factors for DSE were general drivers of discontinuation, independently of side-effects, we ran the same analysis for DOR (Table 2, Figure 3). No physiological risk factors were associated with DOR, but instead were all sociocultural. Catholics and Protestants have lower odds of DOR (HR=0.40, CI= [0.27-0.64], p<0.001) than Muslims, but there are no differences between Muslims and Orthodox Christians. Wealth is protective for DOR, with the 20% middle wealthiest (HR=0.67, CI= [0.50-0.91], p=0.011) and 40% wealthiest (HR=0.60, CI= [0.47-0.78], p<0.001) having reduced odds of DOR compared to the 40% least wealthy. Those not in a relationship were at >3 times more at risk of DOR (HR=3.48, CI= [2.65-4.57], p<0.001) compared to those in a relationship. DOR results were not driven by women who DSE in the non-DOR comparison group, as the results were not changed when removing those who DSE from the comparison group.

**Figure 3:**
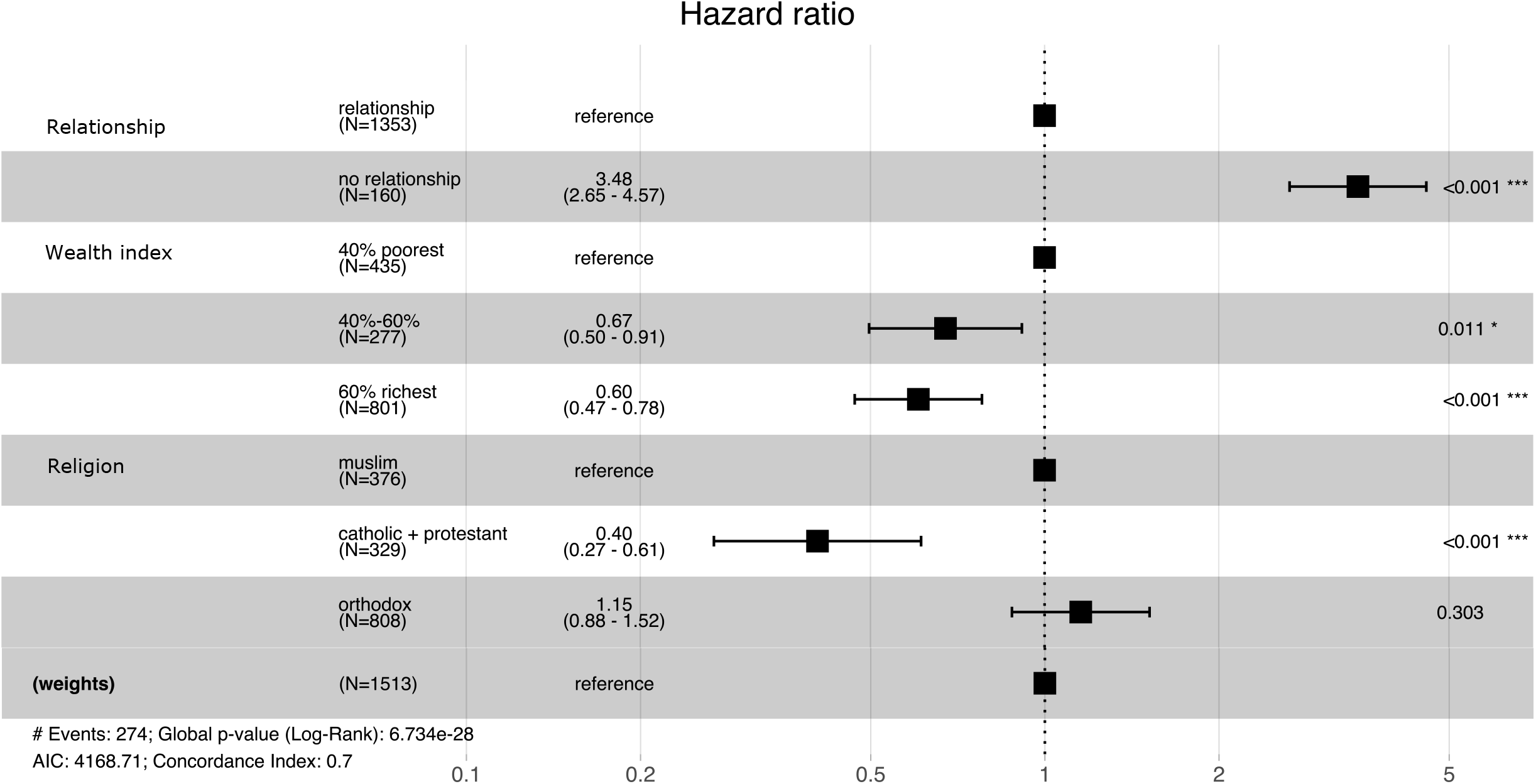
Adjusted hazard ratios and 95% CI for each risk factor for discontinuation due to other reasons (DOR)

## Discussion

Using the Ethiopian 2016 DHS, we investigated whether risk of injectable contraceptive discontinuation due to side-effects (DSE) or other reasons (DOR) was associated with different physiological and sociocultural factors. We found that DSE, but not DOR, was associated with physiological factors likely indicative of low iron status, such as being anaemic, not having received iron supplements and having given birth in the last two years prior to injectable initiation. By contrast, increased risk of DOR was driven not by physiological factors, but instead only by sociocultural characteristics such as lower wealth, not being in a relationship and being of Protestant or Catholic religion. The lack of physiological risk factors for DOR suggests that risk factors for DSE are indeed specific to side-effects driven discontinuation, and not just relevant to discontinuation generally, supporting a potential biological aetiology of DSE. These results have implications for how family planning counselling is carried out and for discourses around the reasons underpinning side-effects.

Anaemia doubles the risk of DSE but has no significant effect on the risk of DOR. This result cannot be explained by the impact of contraceptive use on anaemia: whilst injectable use was associated with a reduced risk of anaemia, and thus women who continue injectable use longer would likely be less anaemic, anaemia was not found to be a risk factor for DOR, despite women who DOR having average shorter episode lengths compared to women who DSE (6.28 vs 7.01 months respectively). This suggests anaemia is a significant risk factor specifically for DSE, rather than discontinuation generally. Previous studies which have found only sociocultural factors to be associated with risk of DSE, may have been confounded by their lack of inclusion of physiological factors, such as anaemia, which are associated with sociocultural factors such as wealth.

Understanding the relationship between anaemia and side-effects in this study is necessarily imprecise due to limitations associated with DHS data. Firstly, the DHS does not collect data on experience of contraceptive side-effects directly. However, it is reasonable to assume that women who DSE experienced more negative side-effects than women who continue use or DOR. Secondly, the contraceptive calendar is measured retrospectively, making it subject to recall bias and post-rationalization. We minimized this bias by restricting the sample to injectable episodes initiated within the last two years. Third, the analysis relies on the assumption that anaemia at interview is a proxy for anaemia at contraceptive initiation. Previous studies^22^ have indeed shown that iron trajectories are fairly stable over time, and the current analysis shows that, if anything, haemoglobin levels increase slightly with injectable use. Therefore, it is reasonable to assume that anaemia levels were either similar or more severe at the time of injectable initiation.

Our other findings also support the role of iron levels *prior* to contraceptive initiation in shaping the risk of DSE. Firstly, having taken iron supplementation with last pregnancy is protective from DSE. Secondly, women who initiated injectable use less than two years after their last birth had higher risk of DSE. High levels of iron are lost during pregnancy, leading to depleted iron stores after birth, particularly so for short interbirth intervals^28^ and for women who did not take iron supplements. Women who have more depleted iron stores after birth may thus experience a greater burden of side-effects. One exception is that women who had given birth 0-2 months before initiating the injectable also had lower risk of DSE than those who had given birth 3-24 months before. This may be due to increased motivation to avoid pregnancy very soon after giving birth. As a proportion of our sample had not given birth in the last 5 years and thus there was not data available for whether they had taken iron supplements with their last pregnancy, we also ran a sensitivity analysis only including women with births in the last 5 years. The effect of iron supplementation remained consistent when analysing these women only. It is also interesting to note that the effects of iron supplementation and time since birth before initiation remain after adjusting for haemoglobin levels, suggesting that depleted iron stores, independent of haemoglobin levels, may themselves also contribute to side-effects. Multiple measures of iron status are thus needed to understand what may be driving side-effects.

Taken together, our findings point towards iron-deficiency anaemia specifically as a driver of DSE. Anaemia may be caused by nutritional deficiencies, pregnancy depletion, or blood loss (iron deficiency anaemia (IDA)) or immune stress causing iron withholding (anaemia of infection (AI)). Our finding that iron supplementation is protective of DSE suggests that iron-deficiency anaemia is particularly important for DSE. If the relationship between anaemia and DSE was driven instead by anaemia of infection, which is characterized by iron withholding and decreased iron absorption, we would not expect to see an effect of iron supplementation on DSE. Future analyses should include biomarkers of immune stress and additional measures of iron status (e.g. ferritin) to understand if anaemia is caused by iron deficiency (low ferritin/low haemoglobin) or iron withholding (normal-high ferritin/low haemoglobin).

Why low iron increases the risk of DSE is unclear. One possibility is that side-effects may present an additional burden on top of the symptoms of anaemia. Another is that there may be a relationship between iron status and reproductive hormones levels. Women with lower natural levels of reproductive hormones have been suggested to experience the most side-effects from typically high contraceptive doses^4^, but whether low iron is associated with lower reproductive hormone levels remains to be further investigated^28^. Studying the response of anaemic and non-anaemic women to different doses of contraception, such as the lower dose injectable, Sayana Press, may present a promising avenue for understanding the interaction of dose, hormone levels, iron and side-effects.

The association between iron status and DSE has potentially important implications for both family planning programs and research. Family planning programs might benefit from providing an integrated service package addressing anaemia as well as supplying hormonal contraception, whereby counselling prior to prescription of contraceptives could add a haemoglobin test. If anaemia is detected, the cause of low haemoglobin could be investigated and treated, either with iron supplementation, in the case of IDA, or relevant antimalarials or antibiotics, in the case of AI. Additionally, the results of this study reveal two gaps in current discontinuation research. Firstly, as discontinuation is a heterogeneous phenomenon which is not currently measured in a nuanced way^2^, studies could benefit from collecting prospective longitudinal data, measuring multiple discontinuation reasons per event, side-effect experiences, and fertility desire measures, which encapsulate fertility planning and motivations within a woman’s context. Secondly, further research on discontinuation and contraceptive development should include additional physiological risk factors, such as iron levels, when examining risk of side-effects. Understanding the causal relationships between physiological factors and side-effects not only may help reduce discontinuation, but also help stimulate research to improve contraceptive technology, doses and service quality for women who continue use, as well as those who discontinue. This utilises a rights-based, quality of care approach^29^ by focusing concurrently on quality of life during use and helping women achieve their fertility goals through either continuation or timely discontinuation of their method of contraception.

## Supporting information

Figure S1: Summary plot of covariate balance before and after conditioning.

Figure S2: Plot showing the number of missing values in each variable in the dataset.

Figure S3: Missing values map showing distribution of missingness across the dataset.

## Data Availability

All data is available upon request from the Demographic and Health Survey website (https://dhsprogram.com/).

## Author contributions

RS and AA developed a proposal and study design for the analysis, with input from EG. RS and BM undertook the literature search. JR did most of the statistical analysis. RS, AA and BM contributed to drafting the manuscript. JR and EG reviewed the manuscript and provided feedback and additions.

## Figure Legends

**Figure S1:** Summary plot of covariate balance before and after conditioning. This plot shows that balance was improved on all variables after adjustment, bringing within the threshold value of .1 for absolute mean differences.

**Figure S2:** Plot showing the number of missing values in each variable in the dataset.

**Figure S3:** Missing values map showing distribution of missingness across the dataset.

