## Supplementary figures and images for "Anaemic women are more at risk of injectable contraceptive discontinuation due to side-effects in Ethiopia"

### Figure S1: Summary plot of covariate balance before and after conditioning.

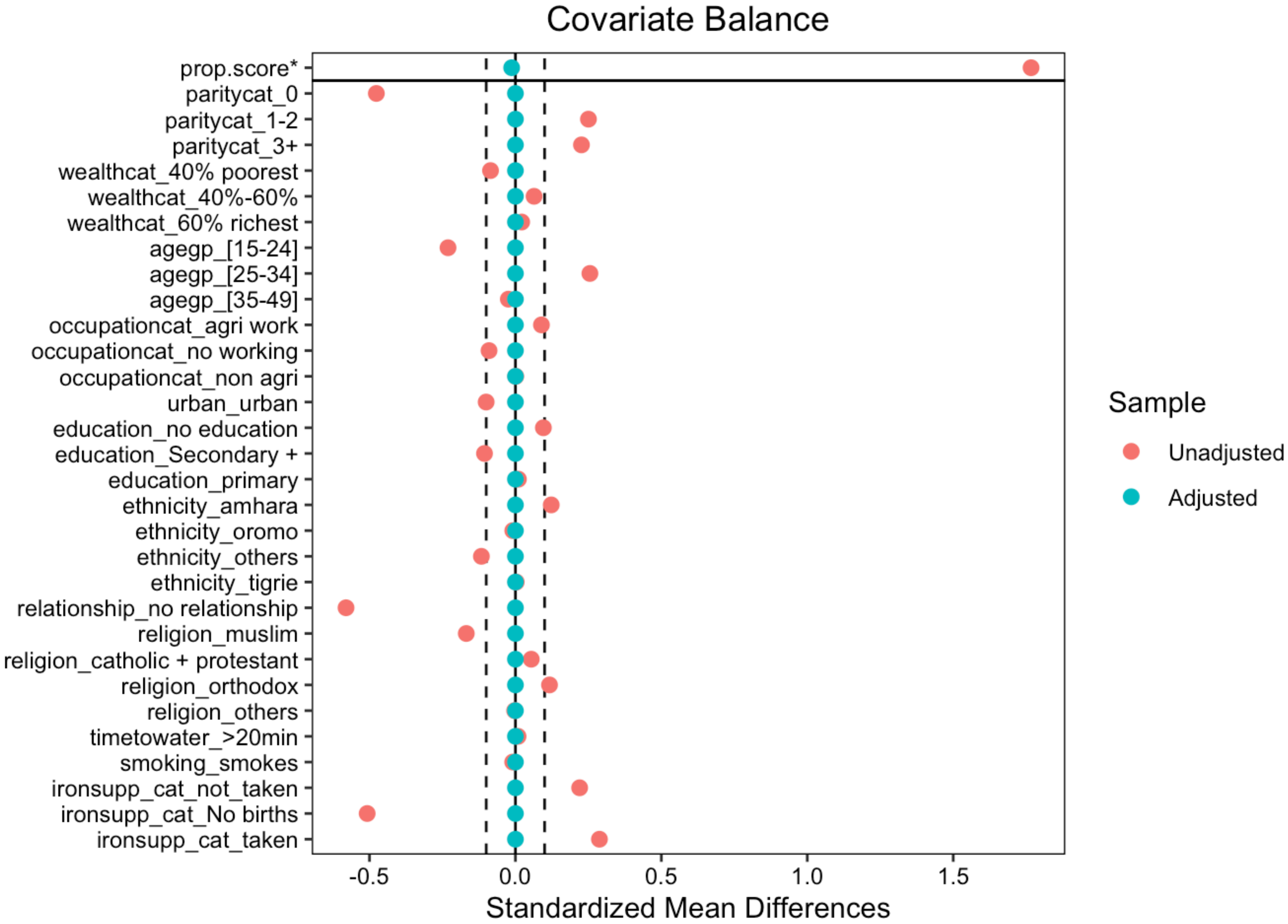

### Figure S2: Plot showing the number of missing values in each variable in the dataset.

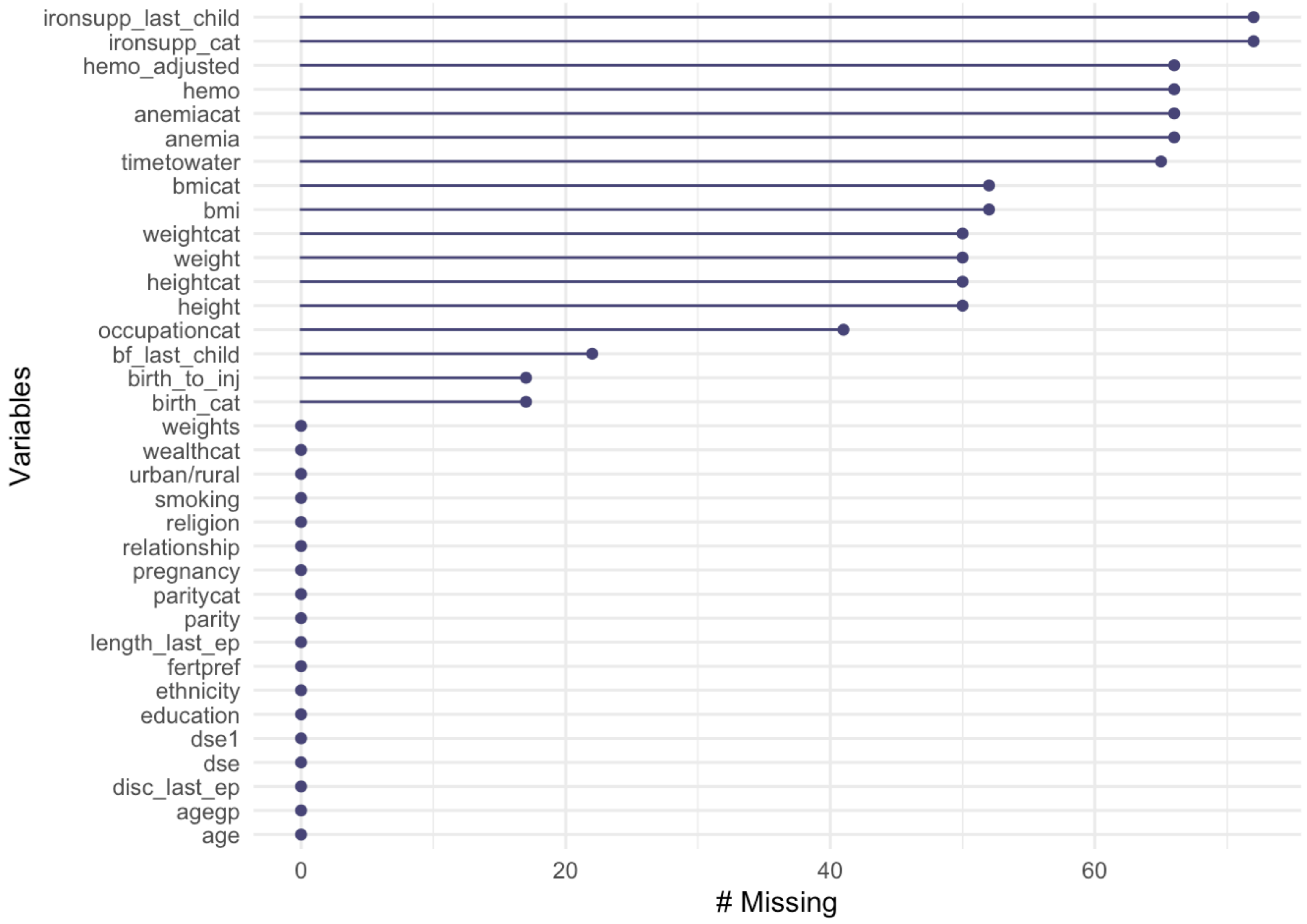

### Figure S3: Missing values map showing distribution of missingness across the dataset.

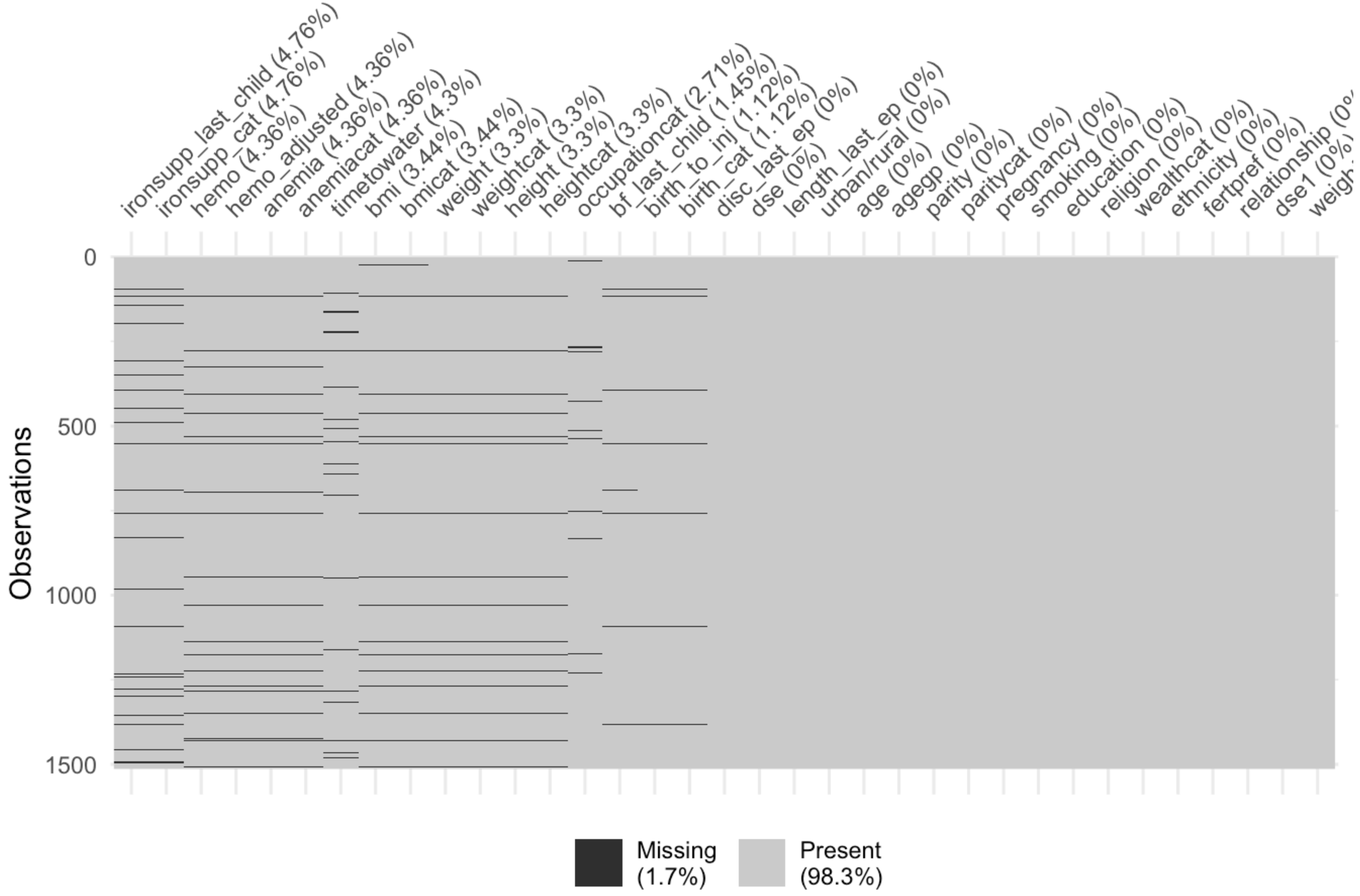
